# Male Allyship to Advance Women’s Global Health Leadership in the Academy

**DOI:** 10.1101/2025.09.17.25335840

**Authors:** Amanda Marr Chung, Ola Alani, Michele Barry

**Affiliations:** Stanford Center for Innovation in Global Health, Stanford University, Palo Alto, California, United States of America

## Abstract

Women are underrepresented in leadership positions within global health. Although women leaders have been shown to foster inclusive work environments and prioritize improvements in women’s health, they face barriers to their advancement, including microaggressions and disproportionate caregiving responsibilities. Male allyship can facilitate the elevation of women into global health leadership roles. This study explores the experiences of global health leaders in academia on male allyship and identifies actions and best practices to support the growth of women’s leadership in global health.

Qualitative semi-structured interviews were conducted with twenty-one global health leaders (11 females, 10 males) from U.S. and Canadian academic institutions. Interviews were recorded, transcribed, and coded utilizing a combined inductive-deductive approach.

Participants identified barriers and outlined potential approaches to support women’s advancement to leadership roles. For the individual male ally, recommendations included completing a self-assessment (to mitigate counterproductive behaviors and biases), engaging in effective mentorship practices, advocating publicly, and serving as a positive role model. Recommendations at the institutional level emphasize the importance of cultivating an enabling environment that facilitates open dialogue, establishing goals and metrics; and implementing allyship training with periodic evaluation. At the societal level, participants suggested promoting early education and shared caregiving to shift cultural norms on gender roles.

This paper provides a framework of actions and resources to cultivate and support male allyship for women’s leadership advancement in global health. Effective male allyship begins with acknowledging power dynamics and an understanding of how intersectionality, beyond gender alone, shapes women’s careers and workplace dynamics. Additionally, mentorship and collaborative peer support are critical to promoting women’s career development. Individual allyship when combined with institutional and societal actions and policies, can facilitate the advancement of women in global health leadership roles.

## Introduction

The world suffers from a severe shortage of women leaders: women make up close to 70% of the global health workforce but only hold 25% of senior leadership positions [1]. Women in the workplace experience macroaggressions in the form of sexual assault and exploitation, gender pay gap, and lack of paid family leave, with the U.S. being one of few countries in the world that do not have national paid maternity leave [2–4]. Microaggressions in professional settings show up when a woman is questioned about her qualifications, not acknowledged for her contributions, and assumed that she is not the leader among a mixed gender group [5,6]. According to the annual Global Health 50/50 report, gender parity has improved since 2018. For example, women CEOs increased from nearly 29% to 35% from 2018 to 2024. However, the world is still far from having an equal number of women as CEOs, senior managers, and board chairs [7].

The political climate has a profound effect on the role of women, which was already fragile at best. For example, in the United States, the dearth of women in leadership positions will likely be exacerbated by the ending of diversity, equity, and inclusion programs in 2025 [8,9]. In 2023, women were removed from high profile university president roles as a reaction to student protests on college campuses [10]. The current U.S. administration will also threaten women’s rights to bodily autonomy, building on the overturning of Roe vs. Wade, which protected women’s rights to abortion for fifty years [11].

Underrepresentation of women leaders and women’s lack of access to supportive networks and sponsors are examples of second-generation gender bias, defined as “powerful but subtle and often invisible barriers for women that arise from cultural assumptions and organizational structures, practices, and patterns of interaction that inadvertently benefit men while putting women at a disadvantage” [12]. In addition to these barriers, mid-career women, especially women of color, tend to be in the sandwich generation, where they have primary caregiving responsibilities for children and aging parents [13,14]. This additional role comes at a time when mid-career women typically need to lean in to get promoted [14,15]. There is a stark disparity between the U.S. and other high-income countries, which have an average of over 18 weeks of paid maternity leave [16]. In the U.S., maternity leave is recognized by workplaces, but it is not always paid [17,18]. Moreover, other conditions specific to women including premenstrual and menopausal symptoms, miscarriages, and abortions merit flexible and paid leave organizational policies, but few provide such accommodations [4,19]. Women pursuing careers in global health often face extended timelines to attain senior positions because these roles typically require fieldwork. This requirement can coincide with the period when many women seek to start families or already have young children [20]. Consequently, these barriers contribute to a phenomenon known as the “leaky pipeline”, in which women are more likely than men to leave academia before securing tenure [21].

### Benefits of women leaders

“Diversity drives innovation – when we limit who can contribute, we in turn limit what problems we can solve.”-Telle Whitney

Historically, women have been excluded from participating in various roles in society including academia, research, and politics [22–24]. Women leaders create more equitable, inclusive, collaborative workplaces and bring different perspectives and approaches than men, including increased empathy and ethical initiatives [25–27]. These approaches have supported climate mitigation and been linked to lower CO2 emissions [28]. Women leaders prioritize the needs of girls and women, vulnerable populations who are often overlooked in policies and research and are more at-risk for diseases such as HIV and malaria [20,29,30]. In research, women principal investigators were shown to increase women participants [31]. In the private sector, gender and racial diversity make good business sense, impacting the bottom line and increasing innovation, sales, revenue, and customers [25,32]. Caregiving responsibilities can make women better leaders, rather than parenthood being perceived as a liability [33–35]. A survey conducted by the Rutgers Center for Women in Business found that respondents doing unpaid caregiving developed and improved workplace skills such as empathy, efficiency, and task prioritization [34].

### Expanding the pool of male allies

As the dominant group, men have a responsibility to use their power and privilege to sponsor women and elevate them to leadership roles. Society suffers from a lack of prominent male allies, often drawing on the same few men to speak out and serve as role models [36]. Moreover, men perceive themselves to be better allies than they actually are [37]. For example, in the 2022 Allyship-in-Action Benchmark Study, 71% of men reported that men “always” or “frequently” give credit to women for their ideas and contributions while only 40% of women agreed with the statement [37]. Men can also support women by stepping up their caregiving, whether it is for children or the elderly. The purpose of this research was to gather concrete actions at the personal, organizational, and societal level that male allies can take to become advocates of women as leaders.

## Methods

### Study Design

The research team utilized semi-structured interviews and developed two interview guides in English, one for each gender (male and female). The guide included the purpose of the study; adapted definitions of male allyship, mentorship, and sponsorship (Table 1); demographic questions; and interview questions [interview guide in S1 Text]. The authors created a list of participants using research team connections and a WomenLift Health list of mid- and senior career academic global health leaders.

**Table 1.**
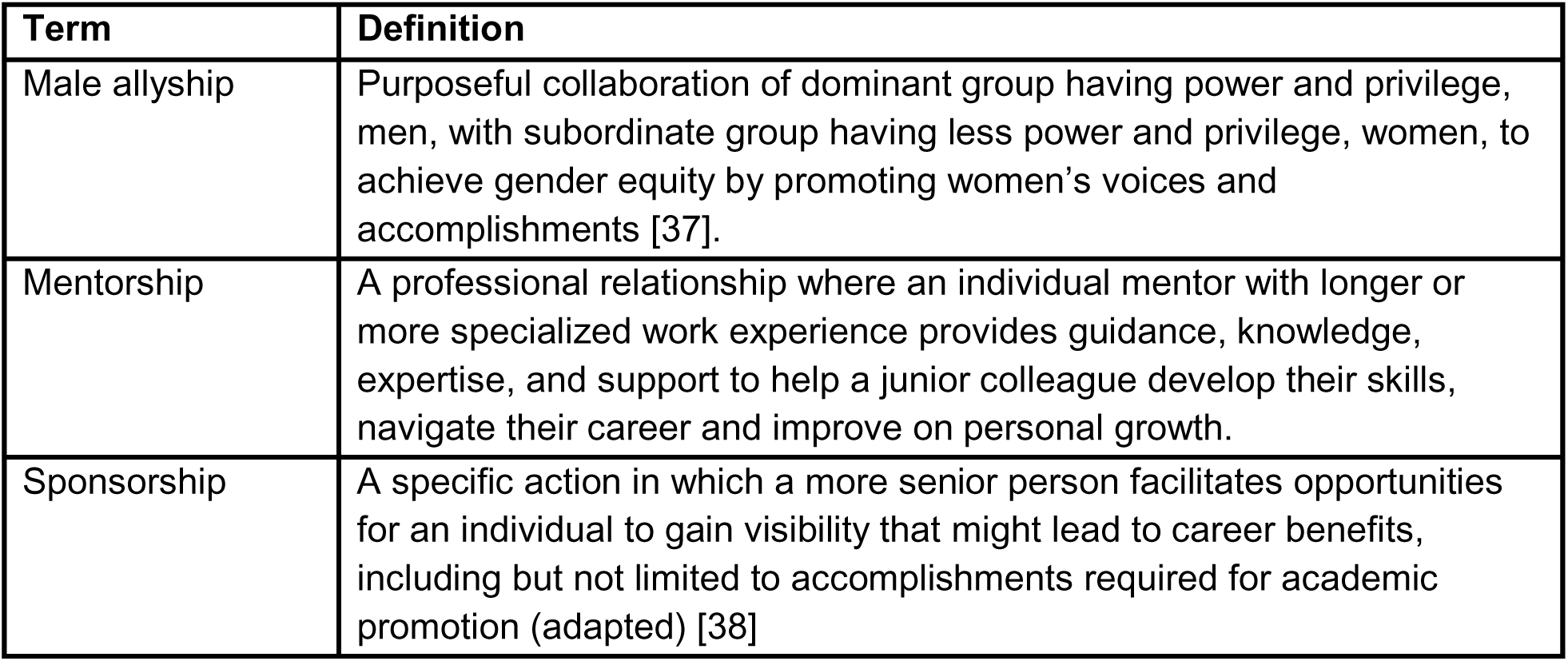
Terms and definitions.

### Data Collection

The study team employed purposive sampling and recruited male and female leaders in global health working in a United States or Canadian academic or clinical institutions between January and November 2024. The team considered the career stage/years of experience, area of study, gender, and ethnicity of participants to collect diverse perspectives. The study team approached participants via email and invited them to participate. The interview guide and written consent forms were shared with the participants prior to the interview. All participants provided their written consent via email, and their verbal consent was recorded. Interviews were virtually conducted in English by the two co-investigators (AMC and OA) via Zoom and captured on an external recorder. On average, interviews lasted 50 minutes.

### Data Analysis

All interview audio recordings were transcribed using Rev AI transcription, and OA reviewed and edited for accuracy. Any identifying information was redacted prior to coding. Data were analyzed by AMC and OA, who have been formally trained in qualitative research. A combination of inductive and deductive coding along with axial and open coding were used. A code book with definitions was created using an individual and collaborative approach. Researchers created an analysis table in Excel utilizing codes to create themes and sub-themes with illustrative quotes.

### Ethical Considerations

This study was approved by the institutional review board at Stanford University, protocol number 72715. All participants received a copy of the consent form via email prior to the interview date and gave verbal consent before starting the interview.

## Results

Forty-seven people were approached: twenty-one agreed to participate; twenty-five did not respond; and one person declined to participate. Eleven participants identified as females and ten as male. Eighteen were from a U.S. institution and three were from Canadian institutions. Thirteen participants identified as white/Caucasian/Jewish, four as South Asian/ Indian/ Indian American, three as black, and one as indigenous. Participants are referred to by coded identifier, gender identity, and five-year age range (e.g., P1, F, 40-44) to preserve anonymity.

Participants spoke about challenges that prevent women from advancing to leadership roles in global health and shared recommendations to mitigate some of the barriers. The study team identified several overlapping themes within the barriers and facilitators and aggregated them based on type of approach and behavior, and whether they were on an individual, institutional, or societal level (Fig 1). Additionally, some participants recommended resources and research such as papers, books, organizations, and checklists. The authors have referenced these where appropriate.

**Figure 1:**
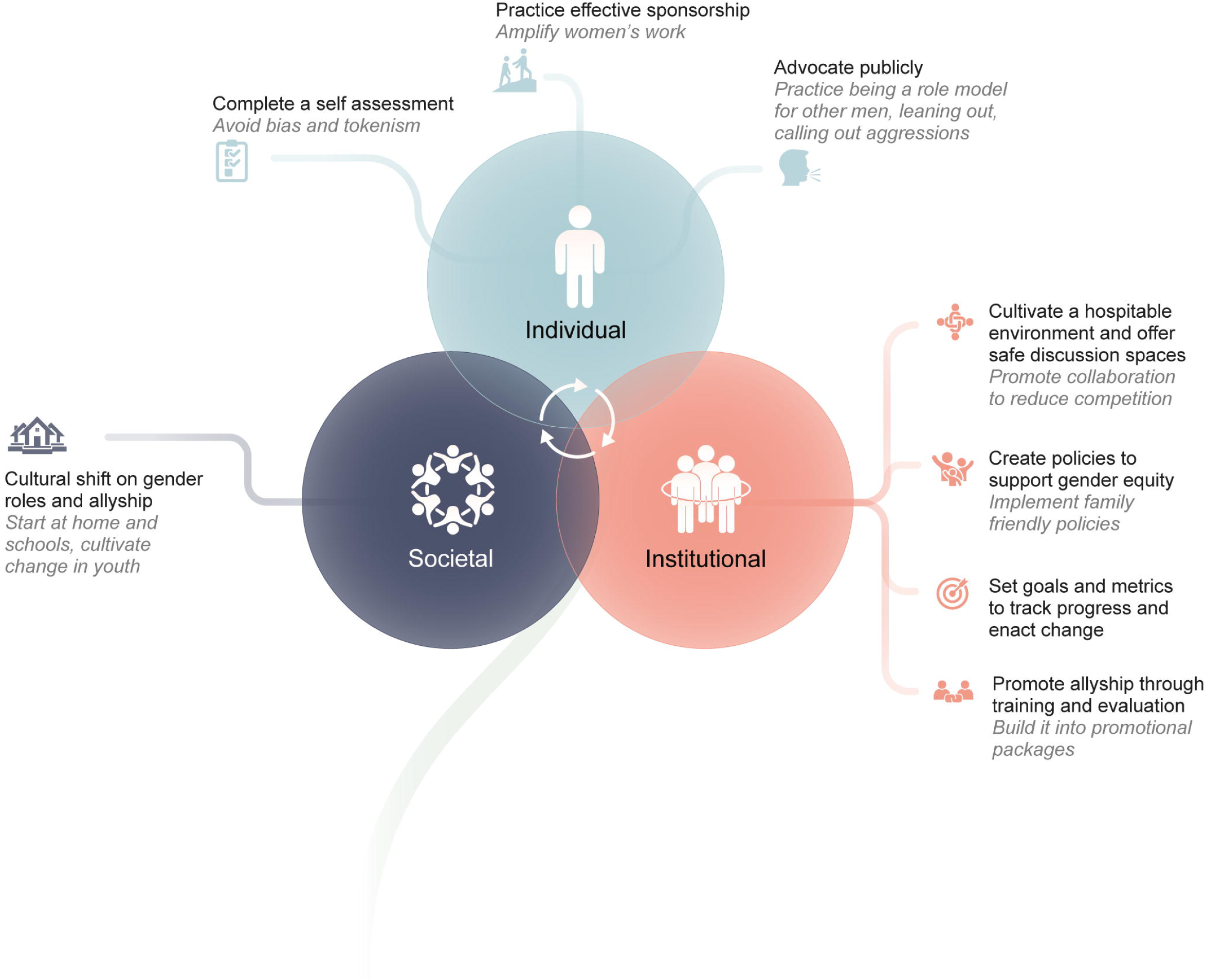
Overview of recommendations for male allyship to promote women’s leadership.

### Allyship

Although the research team defined allyship in the methods, this definition needed further refinement, which they gathered from several interviews. This additional detail included expectations, actions, and behaviors at the individual and organizational levels. According to one respondent:

“Allyship is earned, not something you can self-identify.” -P3, M, 60-64

Individuals who strive to be allies need to do a personal assessment, possess self-reflexivity and intentionality, commit to equity, and recognize and give up power, privilege, and implicit biases. At the organizational level, allyship needs an enabling environment, including promotion by leadership and a culture of embracing diversity.

### Intersectionality

A recurring theme from the interviews was the relationship between intersectionality and allyship. White cisgender, straight men benefit from privilege that also extends but less so to white, cisgender, straight women. When the respondent identified as part of a racial/sexual minority group, they had more awareness of the challenges women in leadership faced and increased solidarity with them. Indigenous mentees in particular felt imposter syndrome. Furthermore, the Me Too movement is a consideration for some but not for all males who identify as straight. For those who are concerned about the misinterpretation of their behavior, they take precautions in their communications and actions, selecting public meeting places and avoiding physical contact and closed door or weekend in-person meetings. Additionally, intersectionality plays a role in finding the right mentor-mentee relationship and is not gender-based.

### Motivation and incentives

Respondents identified several strategies to motivate men to become allies. Intrinsic motivations include being a father to a daughter, personal experience, and the desire to maintain a positive self-image. Additionally, extrinsic incentives, such as recognition and rewards, can encourage men to engage in allyship.

### Individual Actions

#### Self-assessments, reflexivity, and avoiding counterproductive actions

According to respondents, individual actions start with self-assessments related to the power and privilege that males have to relinquish. Once males accept and recognize this, it is their responsibility to lean out, create space, and sponsor underrepresented groups, especially women of color from the Global South.

In order to avoid counterproductive and ineffective efforts, males can:

- Discourage women from taking glass cliff positions (promotions and high-risk leadership positions that are likely to fail [39]).
- Highlight women’s technical expertise in a public facing role.
- Give leadership opportunities with decision-making power to women, providing guidance and support.

Related to the last point, one respondent noted:

“…women in those roles end up taking on more and more roles and actually sometimes to the detriment of their career. So it seemed to be a leadership opportunity, but honestly, it’s a service opportunity… I think the flip side of that is that also takes time away from one’s ability to focus on the things you have to do to get promoted, get more research funding, write more papers, and so on. So I’ve seen…it looks good because women are in those leadership roles, but actually if you kind of dig beneath the surface, what happens is they take on those roles, and I think it just adds to their plate. There’s nothing that comes off their plate in order to play those roles, and that’s a problem.” -P6, F, 50-54

#### Awareness, role modeling, and accommodating caregiving

According to some respondents, men need to be aware of how they are extending offers to socialize after working hours and the type of activities they suggest that may prohibit women from participating due to caregiving responsibilities. Some participants pointed out that men can also step up their caregiving duties by modeling this at work. They can lead by example by making it known that they are leaving early to pick up their children or taking time off to care for their sick child or aging parent. This also extends to mentoring and supervising in advising mentees and employees on work-life integration.

“When she had her first child, she decided to go 80%… She said, ‘… I want to come home to be with the kids’. And I said, ‘Well, so do I. So do all the men that I know, and you don’t just cut your pay 20%…because you feel guilty that if you do go home in the middle of the day or you go to a sports event, a soccer game… there’s not a man I’ve ever met that feels that way.’” -P1, M, 70-74

Men should know at what stage their female mentees/ employees are at in their lives and recognize the impact of caregiving on their work. Men can support women they supervise by allowing them to prioritize their families, accommodating caregiving responsibilities by giving time and grace periods for tasks and projects, especially during difficult periods or crises. An added benefit of caregiving is that it can be a facilitator to connect with employees, students, and mentees.

“I just made some rules for my whole team, and the first rule is family first…children and pets are welcome on all of our virtual platforms… equity is not saying that everybody has to adhere to the same schedule. Equity is saying, we’re going to help everybody succeed. And the people who are in the middle of childbearing, we get that, we care, and we are going to be supportive.” -P8, M, 65-69

#### Mentoring, advising, and sponsorship

Participants also expanded on the definition of mentorship that the study team provided. Some respondents emphasized the importance of having personal relationships with their mentors/mentees and being a role model. A checklist was compiled from suggestions by the respondents (Table 2), including these statements:

**Table 2.**
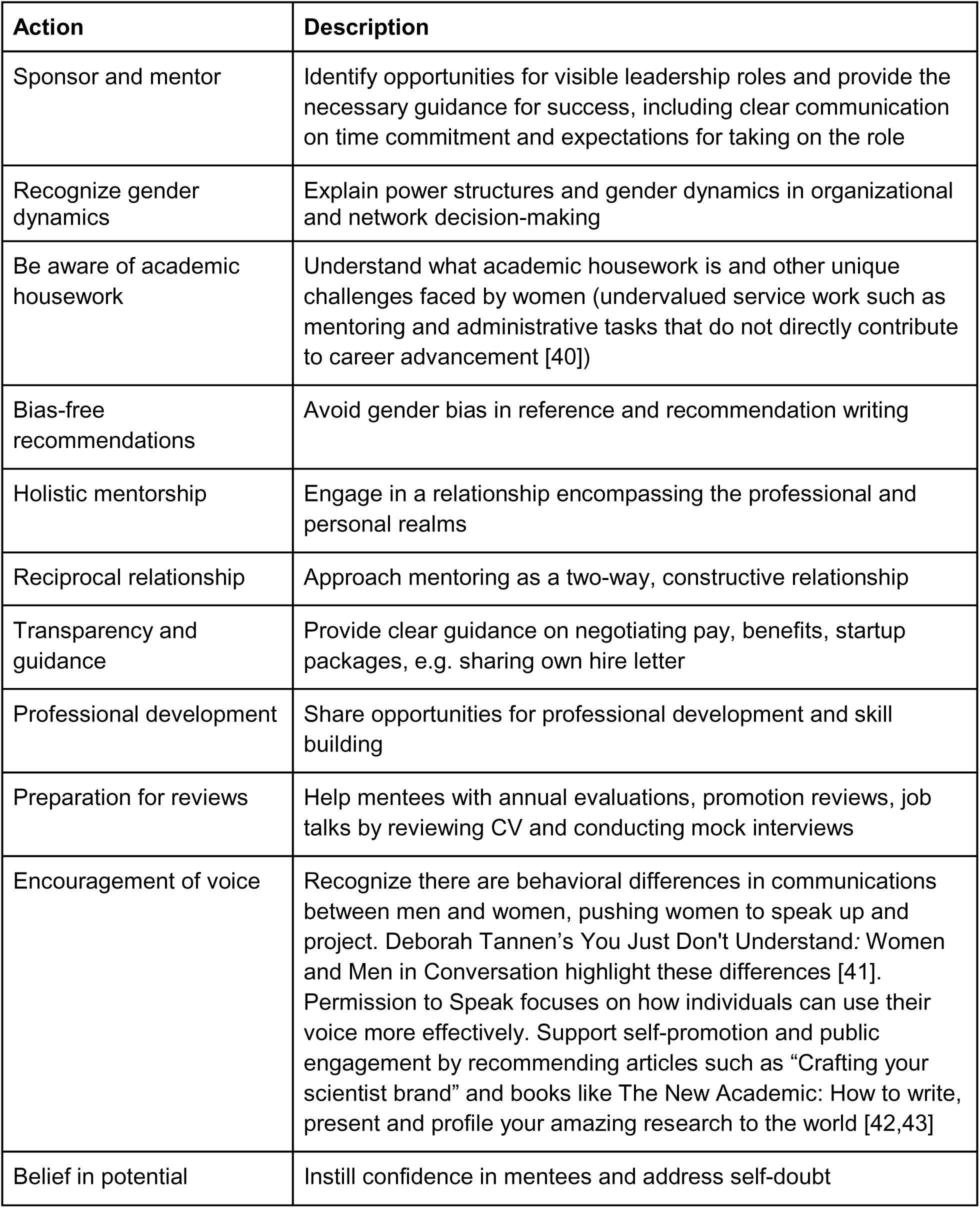
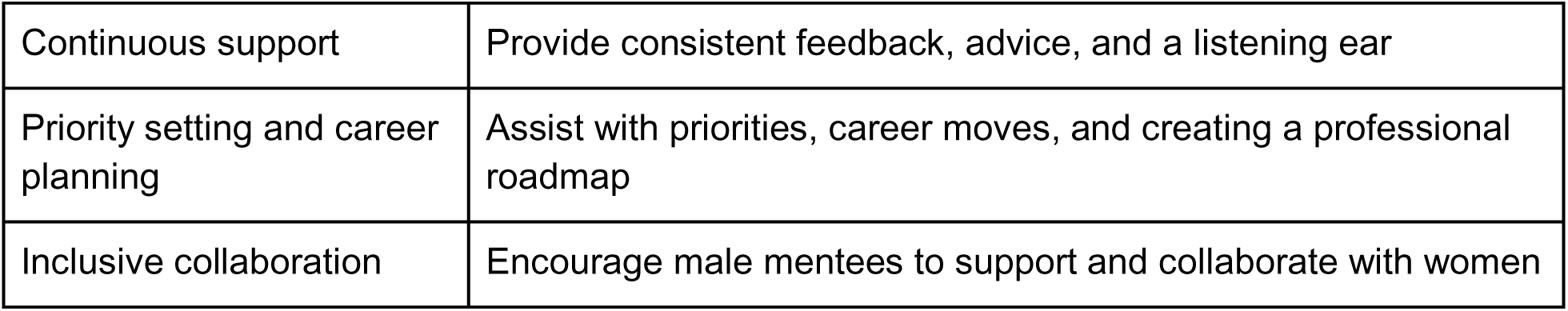
Checklist of actions that make good mentors for women.

| Action | Description |
| --- | --- |
| Sponsor and mentor | Identify opportunities for visible leadership roles and provide the necessary guidance for success, including clear communication on time commitment and expectations for taking on the role |
| Recognize gender dynamics | Explain power structures and gender dynamics in organizational and network decision-making |
| Be aware of academic housework | Understand what academic housework is and other unique challenges faced by women (undervalued service work such as mentoring and administrative tasks that do not directly contribute to career advancement [40]) |
| Bias-free recommendations | Avoid gender bias in reference and recommendation writing |
| Holistic mentorship | Engage in a relationship encompassing the professional and personal realms |
| Reciprocal relationship | Approach mentoring as a two-way, constructive relationship |
| Transparency and guidance | Provide clear guidance on negotiating pay, benefits, startup packages, e.g. sharing own hire letter |
| Professional development | Share opportunities for professional development and skill building |
| Preparation for reviews | Help mentees with annual evaluations, promotion reviews, job talks by reviewing CV and conducting mock interviews |
| Encouragement of voice | Recognize there are behavioral differences in communications between men and women, pushing women to speak up and project. Deborah Tannen's *You Just Don't Understand: Women and Men in Conversation* highlight these differences [41]. *Permission to Speak* focuses on how individuals can use their voice more effectively. Support self-promotion and public engagement by recommending articles such as "Crafting your scientist brand" and books like *The New Academic: How to write, present and profile your amazing research to the world* [42,43] |
| Belief in potential | Instill confidence in mentees and address self-doubt |
| Continuous support | Provide consistent feedback, advice, and a listening ear |
| Priority setting and career planning | Assist with priorities, career moves, and creating a professional roadmap |
| Inclusive collaboration | Encourage male mentees to support and collaborate with women |

“I want my mentees to feel comfortable asking things of me, and I want to feel comfortable asking things of my mentees. And even more importantly, we’ve got to feel comfortable with reciprocal constructive criticism. Otherwise, you’re not a mentor, you’re just a supporter. And that I think is the hardest thing nowadays, to be honest, is this issue of constructive criticism.” - P3, M, 60-64

“Women are just so much less likely to promote themselves, I mean it’s all out there, right? It is nothing new, but when you read an application from a woman, they’ll underplay everything they’ve done. And when you read an application from most, many men, not all men, they exaggerate what they’ve done enormously.” -P4, F, 65-69

“And also I think communication styles tend frequently to be pretty different from men and women. There’s a wonderful book… It’s about how men and women talk and how they hear language. And I think better understanding on both sides about those differences in communication styles would go a long way because until we can communicate effectively with each other, it’s going to be hard to get through this stuff.”-P14, F, 65-69

#### Leaning out and sponsorship

Participants spoke about men in senior positions who hold on to their positions tightly. Instead, they should support their junior colleagues by stepping down and elevating women in their place.

“…I would say older white men in particular, have serious issues with stepping down. They’re so used to power for decades. It really, really threatens them to kind of leave. And so that’s why they hang on for a long, long time. Well after their time is up.” -P1, M, 70-74

Participants suggested a number of ways that men can push women forward while taking a step back, including: seeking women as collaborators on grants and as first or last authors on papers, offering women opportunities to speak on panels, nominating them for leadership positions and awards, creating positions for women or elevating them as co-leaders, giving them airtime, explicitly acknowledging their leadership and skills, and supporting their work and their journey as leaders. Respondents commented that men should have gender equity as a personal criterion when recommending and nominating others while also thinking of Global South representation.

“And one of the reasons I decided to leave and do this kind of later career change is precisely because I did not want to be one of those people who didn’t get out of the way. And the junior folks that I had mentored, it was clear to me a number of them were getting up to full professor.

They were ready to lead, and as long as I was there, there was never going to be that possibility” -P8, M, 65-69

Another respondent also spoke about the importance of junior men being inclusive and collaborative with women:

“And the more we can talk about collective responsibility and saying that it’s important to have diverse teams and that men are also sidestepping … And what I mean by that is that men are willing to say, “look, I’m going to be the second person, the second author or the supporting author.”…Like “I’m comfortable being the deputy, I’m just comfortable being assistant, I’m comfortable doing the secretary role,” because too often, even at that early stages of career progression, men are stepping over women and they may not realize it…That means even any opportunity, whether it is to publish first, whether it is to lead a working group, even the closed door invitations to be invited to a particular meeting or discussion, thinking about how young men can make sure to bring another young woman or another underrepresented person with them into all of these kind of opportunities.” -P7, F, 40-44

#### Public actions

##### Meetings and panels

Respondents agreed that panels should be intentionally designed with gender equity in mind. They recommended that organizers, speakers and panelists, specifically men, should review meeting and conference panel invitation lists to ensure gender representation. Overwhelmingly participants thought men in leadership need to be vocal about not participating in manels (an all-male panel). A couple respondents referenced public statements by prominent global health leaders pledging not to be on a manel [44]. However, one participant spoke about understanding the audience and context before completely dismissing being on a manel, e.g., the only opportunity to access a certain audience. Other participants highlighted the importance of bringing on qualified speakers to avoid tokenism, as illustrated by this quote:

“So I think for example, when you’re putting together a program…a seminar…a [disease X] conference, it’s important to pay attention to gender equity, but it’s really important to…make sure that …there are leading women scientists …and not as an afterthought. Because nobody wants to be put in a position where it’s obvious that they’ve been added for some kind of diversity to something that isn’t really their area of expertise. It isn’t fair, and it sets people up for not good outcomes. So I think tokenism is a challenge and it’s important to really avoid that.” - P8, M, 65-69

##### Bystander Interventions

Participants described actions by allies to call out sexism, inequitable treatment and behavior, and microaggressions. Those actions included redirecting the conversation or question, speaking up for other women, intervening either one on one or in a group, pointing out inappropriate communication or actions by men about women, and deflecting where appropriate. Additionally, participants spoke about the need for meeting facilitators and panel moderators to call out men for their sexist behavior in the moment or follow up privately afterward. Some participants mentioned that there are certain facilitators to intervening on microaggressions such as intersectionality and not being part of the dominant group. The last sentiment was expressed in the following comment:

“A lot of it [decision to intervene] has to do with what you perceive as the intentionality of things, because anytime you correct someone publicly, it’s pretty humiliating for them…But if it’s somebody that’s being arrogant and really inappropriate and you don’t want this to be perceived as accepted, and you do have to step in the moment…I’ve stepped in right in the moment and

…I’ve gone afterwards and said, “She’s a doctor too. So if you’re going to do doctor for everyone, you better do doctor for her as well.” But I didn’t say it in the moment because I thought this was a person that could learn from that …and I didn’t want them to feel like I was making them look bad in front of everyone else.” -P15, M, 55-59

“I’ve got one moment in recent history in Africa where I was sitting with a junior researcher who was a woman and had two bosses who were male…sitting around her kind of joking about what she should be doing and what a woman’s sexual role is. And being able to say, ‘well, in my work you wouldn’t be able to talk about it like this. And I’m sure it makes her feel very uncomfortable.’” -P9, M, 50-54

### Institutional/Organizational

Some respondents indicated that rather than relying on individual actions, institutions should take the lead in enacting change to ensure gender equity in the workplace. These actions can include creating a hospitable workplace environment and supportive policies, developing goals and metrics, and requiring regular training and evaluations.

“But generally, I think it to be hard for men to make room for women without some institutional push, institutional rewards…So I think the institutions that have created this workspace have more responsibility than the men themselves…They have more responsibility to institutionalize, making room for women so that men do not have the option.” -P10, F, 45-49

#### Workplace environment

##### Workplace culture

All respondents spoke about workplace culture and the impact on researchers and their career advancement, with several emphasizing that the impacts and solutions are not exclusive to women. Some respondents acknowledged that academic environments breed competition rather than collaboration:

“I think what prevents men is just they’re focused on their own career trajectories and just trying to succeed and survive in academia themselves. So I think that pursuit of productivity and focus on climbing the academic ladder is probably what prevents them most from thinking about making space and pulling female colleagues along during that process…it is a struggle at our one institution to keep one self-funded and keep productive enough to get those promotion hoops behind you…it’s hard when you’re in the thick of that to think about making space for others.” -P6, F, 50-54

To address the competitive environment, fostering a hospitable workplace culture needs strong leadership who will recognize disparities, advocate for equity, act on any instances of workplace harassment and aggression, and set up checks and balances. Institutions should actively recruit leaders who value diversity and gender equity. Several participants suggested a reframing from a zero-sum game to a rising tide will lift all boats mentality, providing concrete recommendations that institutions could accomplish this by promoting allyship, collective benefits, and engagement by men with women. Institutions can also create spaces for success, promoting a collaborative environment, and a work culture that discourages competition and values everyone’s contributions. According to one participant:

“For NIH dollars, there’s only so much to go around. You’re in a cardiology or a nephrology program doing research, and so are your women peers. And it is a more competitive program. And if you do succeed and get tenure… it’s easy to be less competitive because you have the protections…It’s really on institutions to structure it such that…there isn’t some kind of winnowing. You should take on the people you think you need to achieve your mission. And you should try and ensure that they all succeed. And if everybody knows that, that it’s a level playing field and it’s not, some kind of hunger games struggle, it’s better for everyone. And that departments have to do that, chairs have to do that, deans have to do that.” -P8, M, 65-69

Additionally, some respondents spoke about the impact of having a collaborative leadership model in reducing competition and supporting women’s advancement where a leadership position is shared between two individuals with different responsibilities.

“I don’t know why we had to have a single director…it’s so much more pleasurable to do work in teams and to lead as team leaders. Maybe you have to divide up to certain responsibilities by skills if you have to, but boy, if we just created more environments where it’s not always about competing with your peers, but really working with more than one person to lead…it’s just more fun and it’s more effective and it’s more sustainable because nobody’s doing their leadership role as their only job… Institutions should strive to create a legacy of promoting women to senior leadership positions and co-chairing commissions by integrating these actions into their policies and practices.” -P5, F, 45-49

##### Discussion spaces for men

Several respondents recommended providing men with opportunities to actively and regularly engage in the gender equity conversation. Participants talked about the need for dedicated safe discussion spaces specifically for men outside of women’s conferences and that these should be skillfully structured and facilitated conversations. The spaces should allow for men to voice concerns, ask questions, and exchange knowledge. Men should be able to discuss gender equity, commitment, and accountability. Those spaces could include learning about the differences in communication styles between men and women and addressing related issues. Such spaces could have facilitated discussion relying on case studies and work towards action.

“…I feel like there will be a lot of men who might be willing to lean out or to be allies, if they have a safe space to have these conversations with women without it being accusatory or pointing their fingers or anything like that… I would think, but part of the challenge for them is that they don’t even know how women feel or where women are coming from…whenever we are trying to advance women’s issues, we tend to focus on women and invite women, and men are not always there…We need to try to attract them to come in, if they think it’s safe to come and to say what they feel. If they have to admit their ignorance, it needs to be a safe space for them to admit their ignorance, but if they don’t feel like it’s a safe space, they may not, even if inside they want to.” -P12, F, 45-49

##### Creating workplace peer-to-peer solidarity

Several respondents emphasized the importance of peer-to-peer support to uplift women and reduce competition. Some respondents pointed to the mutual benefits of fostering interpersonal relationships between men and women. Respondents provided recommendations for men to support women colleagues. For some participants, these relationships may arise informally, but institutions could facilitate this through the creation of mixed gender peer mentorship groups or project groups where individuals alternate between taking the lead on a project and providing support. Some recommendations by respondents on actions by male peers include:

- Attending women’s talks, giving encouragement and providing feedback at the conclusion
- Reminding women to take credit and acknowledging women publicly for their contributions
- Collaborating on grants
- Transferring legacy/projects to a woman when leaving/retiring from an institution
- Creating career advancement opportunities for women
- Providing peer mentorship
- Sharing successful approaches for public recognition
- Discussing how to navigate career challenges
- Recognizing when a female colleague is overwhelmed or handling competing priorities and offering support

One respondent spoke about the valuable support she received from male peers:

“I think about allyship really specifically in terms of thinking about someone kind of promoting you or supporting you or sponsoring you, and also just maybe giving up some of what they have to move things forward. I really haven’t had that a ton except actually in the peer spaces. So I have to say that honestly it’s peer male colleagues similar generationally, not by age, but generations in terms of when we came in together, what kind of work we’ve been doing together when we’ve been on the same teams that have really helped.” -P5, F, 45-49

##### Meetings, Conference Panels, Commissions, Committees

Some respondents provided tools and recommendations for institutions and groups to ensure equitable representation in meetings and conferences. In order to support equal representation for women, institutions can incorporate a check for gender balance into standard operating procedures when developing invitation lists to high-profile meetings and make space in agendas for women’s perspectives, including providing speaking opportunities for underrepresented women.

These suggestions were illustrated by this quote:

“…let’s say we are organizing a new conference…a new commission or a panel… right at the start, you’ve got to say, this conference will be co-chaired by a male and a female. This panel will be co-moderated. And then at the end of this initial round of selection, have we made sure (of)… at least gender parity? Sometimes it needs to be more than gender parity…Sometimes it needs to be an all women commission because that’s what the topic requires. So having that intentionality I think is critical.” -P2, M, 50-54

#### Policies

Participants discussed the responsibility of institutions to recognize gender disparities, including biological differences that impact women, and enact cultural change by developing policies and guidelines that work towards equity. Policies are necessary to support the sexual and reproductive health and rights of women in the workplace throughout the lifecourse, from menstruation, family planning, childbearing, menopause, and domestic violence.

Participants provided several examples of improvements needed in institutional policies (Fig 2).

**Figure 2.**
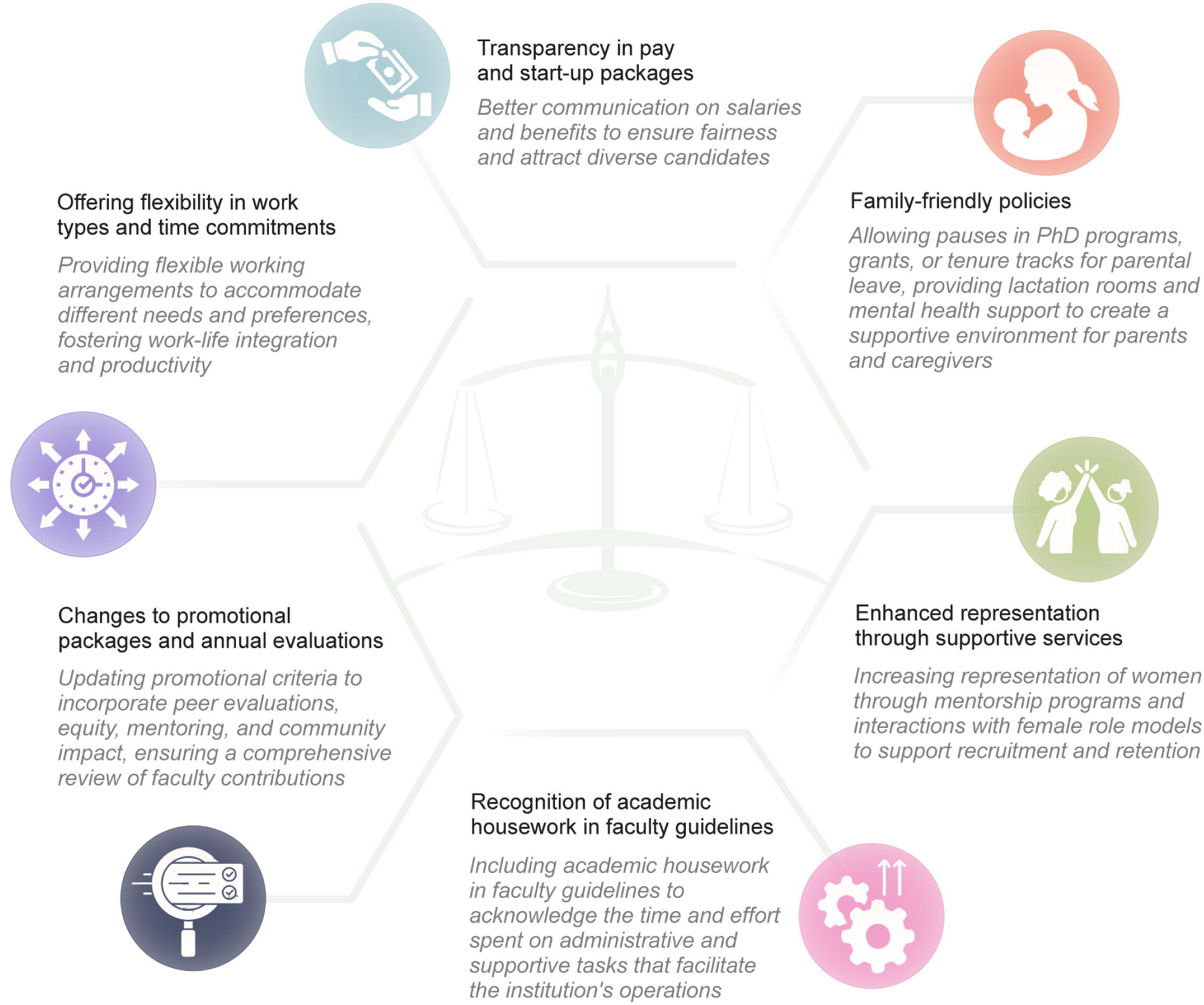
Recommendations for institutional policies to promote gender equity.

“There’s some things in the department I’m just cringing…not allowing women to work part-time. That’s just ridiculous. Many of them obviously have children. They could do well by working part-time, and they have these rules in place that they won’t take part-time faculty, when I was chief of medicine…we pushed for letting women work and having flexible work hours and having a covering system for each other…So I think that’s kind of a structural change that I could see that we need to advocate for, that we have advocated for.” -P21, M, 60-64

Another respondent suggested that the way to stimulate institutions to promote gender equity in leadership positions was through funder policies:

“If you were going to capture a group that was going to be able to incentivize more gender equity related to leadership, really it’s the funders who could be asked to be looking at what’s the proportion of leadership from different genders.…But I think if you’re wanting to motivate organizations and institutions to sort of have a more gender balanced leadership, then it seems like the money is going to be the thing to drive it. Otherwise, you’re just depending on the institution, so of their own goodwill decide that this is a priority, which would be nice.” -P16, F, 45-49

#### Goals and metrics

Some respondents suggested developing organizational goals to achieve gender equity and then creating metrics on gender parity and salary gaps for accountability and tracking of progress. Alternatively, institutions can devise a scorecard with red, yellow, and green to measure performance and would need to regularly re-examine and revise it. These goals and metrics, when paired with interventions and communications, are an effective tool to monitor gender equity and adapt course if needed. They should inform hiring, promotions, and annual reviews. One respondent suggested a town hall to review metrics and processes that are in place to allow for greater transparency, accountability, and improvement.

#### Professional development and evaluation

Respondents discussed the need for required training and self-assessments to raise awareness of and educate faculty on gender disparities and bias, promote equity, and foster male allyship. Some respondents recommended these topics to be part of leadership training and use positive and inclusive language to engage men effectively. Areas of focus could include understanding biological differences and potential work accommodations, cultural expectations, structural barriers faced by women, teamwork, and mentorship. To encourage allyship, the training could provide a toolkit for men that might include checklists and real-life case studies.

Several respondents recommended that a commitment of allyship be built into promotion packages, salary reviews, or annual evaluations. To gain a full understanding of an individual’s allyship these evaluations could include feedback from peers and mentees with recommendations on how to help faculty improve their practices. Based on this feedback, institutions could then recommend trainings and professional development to encourage allyship. Another spoke of a ‘citizenship credit’ which are extra points faculty can get for non-monetary work (e.g., attending a workshop on gender equity).

### Societal

Participants spoke about changing cultural and societal norms by fostering male allyship at an early age. This could be introduced into formal education at the primary and secondary school levels, with discussions on power, privilege, gender, intersectionality, and gender stereotypes and biases. Some respondents pointed to the importance of families also discussing caregiving, pay equity, and gender norms. Several respondents mentioned the support from male life partners who offered advice, shared responsibilities, and advocated for them.

“I again emphasize more and more that we can [*introduce*] the discrepancies of power as early as we can in education, and if we can get it into primary and secondary education as part of formative education, and then whenever formal education starts for those that enter university and all global health programs should be having a mandatory course on power and privilege and using intersectionality and gender. It should not be an add-on course essential. That is a required aspect of formative education. I think we’re going to keep running into those that don’t see how they are the enablers, the bystanders, or actually those that are propagating our power imbalances. -P7, F, 40-44

Similar to the recommendation for academic institutions to offer safe spaces for men to discuss gender, schools and community organizations might organize boys and men’s groups where case studies or examples from girls and women could be presented and discussed. According to one respondent:

“But I hear time and time again, even from our elders in indigenous contexts, that… we don’t have enough focus on men and men are losing their way, and it perpetuates these kinds of toxic behaviors because they really are not getting the support in these kinds of spaces that women have been for many years in some… And without giving them spaces to help them find their way back, it’s going to be really hard to have them be good participants of community and recreate the matriarchal societies that we had prior to colonization because of the inputs from colonial societies on these patriarchal norms.” -P20, F, 40-44

Another participant spoke about how ingrained it is in our culture for women to do the caregiving and the difficulties of juggling these responsibilities with a career in global health:

“I pay attention more on how life is affecting women differently than men, especially in global health settings where women have that dual responsibility of showing up as professionals at work, but they’re also like their primary caretakers at home and in charge of their kids and their families… Women definitely have disproportionate roles outside of the work environment.” -P12, F, 45-49

## Discussion

The purpose of this study was to understand the experiences and perspectives of global health leaders on male allyship and present actions and best practices to support the advancement of women leaders in global health. Not all recommendations from the participants fit neatly into one level, whether individual, institutional, or societal (Fig 1 shows the interconnectedness of the different levels). For example, mentoring is an area where traction can be made at the individual level with more training while also being scaffolded by institutional policies and metrics to support elevating women into visible leadership roles. Women face unique challenges related to biological differences as well as societal and cultural expectations, especially around caregiving.

Both institutions and society will benefit from actions that males take to create an academic environment that discourages a zero-sum game mentality, elevates women to leadership positions, and reduces gender disparities. Changing societal norms about shared caregiving to emphasize the benefits of men as caregivers can lead to healthier families and relationships [45–47]. Greater gender equity can also bolster economies and innovation, leading to shared prosperity [48]. National policy changes such as setting quotas for female political representatives has increased legislative seats for women in Mexico, Nicaragua, and Rwanda, while establishing board representation mandates has resulted in more women’s board seats in France and Norway [49].

Our research aligns with findings that the degree to which individuals identify as male allies can be influenced by intersectionality. Participants are more likely to empathize with the challenges women face in academia if they have experienced similar challenges as a member of an underrepresented group. Prior research has shown that marginalized groups find solidarity and support through shared experiences [50]. Peer-to-peer support and collaboration were postulated as an effective allyship method that can improve work environments and career pathways for all genders [51–53]. This needs to happen in tandem with a shift in institutional policies and rewards systems in academia. Tiokhin et al. argues for placing a greater emphasis on group contributions rather than just individual [54]. Participants spoke about allyship as a continuous practice of shifting attitudes and actions that works towards uplifting women and paving the way for them. Recommendations offered by participants for better allyship (including mentorship) have been echoed by research reviews and insights across various academic disciplines and fields [20,55–58]. Sinha et al. offers ten strategies for male allyship to support women in academic medicine that include individual, institutional, communication, and inclusion-based approaches (S2 Table has select resources and tools offered by participants and the research team to support male allies) [59].

### Limitations

The study was restricted to mid and late career male and female global health leaders who were based at academic institutions in the United States and Canada. Although the team selected participants who represented a diversity of perspectives, they did not interview participants who represented some ethnicities, including those who identified as Middle Eastern, Southeast Asian, East Asian, and Latin American, nonbinary, or transgender. The team was also unable to interview any participants based at a historically black college or university. For several interviews, the participants had limited time, which prevented the researchers from further probing.

#### Next steps

Further research is needed to corroborate the findings of this study in other geographies. The team plans to gather feedback from mid-career African physicians on their recommendations to adapt the resources, tools, and recommendations. At the same time, there is a need for public advocacy, which can be achieved through media engagement and reform of institutional policies to implement these recommendations effectively. Expanding engagement across various regions can foster male allyship and contribute to the establishment of a global male allyship movement. While this study focused on male allyship, achieving gender parity will also require allyship by women and prioritizing the elevation of diverse leaders from underrepresented groups, such as transgender and indigenous women. The global health community needs to not only ensure we have more women leaders but that they also embody underrepresented minorities and are nationals of low- and middle-income countries, especially as the field seeks to establish more equitable global health partnerships. The authors hope this piece inspires men to take action and join others to create a movement that will advance the field of global health and beyond.

## Supporting information

Supplemental Text 1. Interview questions

Supplemental Table 1. Resources and tools to support allyship

## Data Availability

The data set is available upon request from the corresponding author.

## Acknowledgments

The authors extend their gratitude to the research participants for sharing their experiences and insights. They also acknowledge the valuable contributions of Dr. Nicholas Zehner, Dr. Madhukar Pai, Rachel Knopf Shey, Dr. Magali Fassiotto, Dr. Mollyann Brodie, and Dr. Lisa Rogo-Gupta, who provided feedback on the qualitative instrument and offered important resources.

## Supporting information

**S1 Text. Interview questions.**

**S2 Table. Resources and tools to support allyship.**

