## Supplemental Text 1. Interview questions for "Male Allyship to Advance Women’s Global Health Leadership in the Academy"

**WomenLift Health Male Allyship Study**

**Interview Guide for participants (semi-structured one on one interviews)**

**Length:** 45 minutes to 1 hour

**Purpose of survey: C**reate a framework that identifies male allies, mentors and sponsors from the perspective of global health male and female leaders in academic and clinical institutions and generate recommendations and best practices for how those who want to be powerful allies and mentors can help to advance women’s leadership in global health.

Interviews will be conducted via Zoom and taped via a recorder with your permission.

### **Definitions**

You have been identified as an exemplary ally, mentor or sponsor. We appreciate your time to complete this interview. These are some definitions so we can use equivalent language.

- Male allyship:
  - For the sake of this project, which is focused on how to help women achieve gender equity, male allyship is defined as *Purposeful collaboration of the dominant group having power and privilege (men) with subordinate group having less power and privilege (women) to achieve gender equity by promoting women’s voices and accomplishments.*
- Mentorship: *a professional relationship where an individual mentor with longer or more specialized work experience provides guidance, knowledge, expertise and support to help a junior colleague develop their skills, navigate their career and improve on personal growth*.
- Sponsorship: *A specific action in which a more senior person facilitates opportunities for an individual to gain visibility that might lead to career benefits, including but not limited to accomplishments required for academic promotion.*

### **Demographics**

- Age or age range
- Gender identity
- Pronouns
- Current occupation
- Title
- Affiliation
- Academic degrees
- Years of work experience
- Race /ethnicity
- Place of birth
- Global health focus (clinical, research, education)

**Men’s Copy**

### **Open ended questions:** For the purpose of these questions, we will use the terms allies, mentors, and sponsors interchangeably.

1) Why do you think you’ve been nominated as a powerful, effective male ally/mentor/sponsor? What do you think you’ve done that has had others call you that?
2) Do you mentor men differently than women you’ve championed over time? If yes, in what ways do you pursue being a male ally/mentor/sponsor differently for women?
3) How would you advise women to advocate for themselves if they are your mentees?
4) Are there situations in your career where you have seen men intentionally ‘lean out’ to create space for women? What has that looked like?
5) What prevents men from becoming effective allies? Can you offer examples?
6) What examples, if any, of well-intentioned but ineffective or counter-productive male allyship practices come to mind? What advice you would give to men to avoid being counterproductive?
7) Has the **“**me too” movement made you more uncomfortable talking to women or being a male ally? If so, how did it affect your actions towards women colleagues and mentees?
8) Research shows that mid-career men are more competitive with women who are their peers than when men are older and more established. Do you see any way to create more solidarity in peer-to-peer relationships?
9) Have you ever observed male colleagues derailing a woman’s success, harassing, or questioning her qualifications simply because of her gender? Did you get involved in any way? Why or why not?
10) If we created a toolbox of best practices for men in academic institutions to become better allies, what would it contain? These could be at the personal, institutional, or societal level.
11) How would you disseminate the toolbox of best practices?
12) What could incentivize men to share the toolbox with each other?
13) Are there any additional concrete actions that can be taken by male allies, sponsors, and/or mentors to support women leadership in global health and level the playing field?
14) Which global health leaders (of any gender) based in a US or Canadian academic/clinical institutions would you recommend us to interview for this project? We are also interested in male successful mentors in areas other than global health. Please list 3 and their areas of expertise.

**Women’s Copy**

### **Open ended questions:** for the purpose of these questions, we will use the terms allies, mentors and sponsors interchangeably.

1) Why do you think you’ve been nominated as a powerful, effective mentor/sponsor? What do you think you’ve done that has had others call you that?
2) Do you mentor men differently than women you’ve championed over time? If yes, in what ways do you pursue being a mentor/sponsor differently for women?
3) In what way have male allies played a role in your academic success. Please give specific examples?
4) How would you advise women to advocate for themselves if they are your mentees?
5) Are there situations in your career where you have seen men intentionally ‘lean out’ to create space for women? What has that looked like?
6) What prevents men from becoming effective allies? Can you offer examples?
7) What examples, if any, of well-intentioned but ineffective or counter-productive male allyship practices come to mind? What advice you would give to men to avoid being counterproductive?
8) Research shows that mid-career men are more competitive with women who are their peers than when men are older and more established. Do you see any way to create more solidarity in peer-to-peer relationships?
9) Have you ever observed male colleagues derailing a woman’s success, harassing, or questioning her qualifications simply because of her gender? Did you get involved in any way? Why or why not?
10) If we created a toolbox of best practices for men in academic institutions to become better allies, what would it contain? These could be at the personal, institutional, or societal level.
11) How would you disseminate the toolbox of best practices?
12) What could incentivize men to share the toolbox with each other?
13) Are there any additional concrete actions that can be taken by male allies, sponsors, and/or mentors to support women leadership in global health and level the playing field?
14) Which global health leaders (of any gender) based in a US or Canadian academic/clinical institutions would you recommend us to interview for this project? We are also interested in male successful mentors in areas other than global health. Please list 3 and their areas of expertise.
