## Supplemental Table 1. Resources and tools to support allyship for "Male Allyship to Advance Women’s Global Health Leadership in the Academy"

**S2 Table. Resources and tools to support allyship**

| **Level** | **Topic/Action** | **Resource** | **Recommended by (respondent or research team)** | **Published /written by** | **Summary** |
| --- | --- | --- | --- | --- | --- |
| Individual | Self-assessments, reflexivity, and avoiding counterproductive actions | [What Every Health Researcher Needs to Know About Health Equity: Privilege, Oppression and Allyship](https://www.youtube.com/watch?v=yiWZK2AxF7M) [1] | Respondent | The Centre for Healthcare Innovation, University of Manitoba | A recorded session that explores privilege, understanding health equity, and utilizing allyship |
| Individual | Public actions, Meetings and events | [Guidance for Avoiding All Male Panels](https://indonesia.un.org/sites/default/files/2021-06/Guidance_No%20Manel_0.pdf) [2] | Research team | United Nations Indonesia | Recommendations on how to avoid male panels for organizers, panelists, and audience |
| Individual | Mentoring, advising, and sponsorship | [Mentoring Checklists](https://cfe.smhs.gwu.edu/mentoring-checklists) [3] | Research team | The Center for Faculty Excellence, The George Washington School of Medicine and Health Sciences | Mentoring checklists for mentors and mentees for initial and follow-up meetings to ensure a productive, long-term relationship |
|  |  | [Avoiding Gender Bias in Reference Writing](https://csw.arizona.edu/sites/default/files/avoiding_gender_bias_in_letter_of_reference_writing.pdf) [4] | Respondent | Commission of the Status of Women, University of Arizona | Guidance on how to avoid gender bias in reference writing |
|  |  | [Avoid Implicit Gender Bias in Recommendation Letters](https://diversity.ldeo.columbia.edu/sites/default/files/content/Avoid%20gender%20bias%20letters%20Dutt%202019_0.pdf) [5] | Research team | Lamont-Doherty  Earth Observatory, The Earth Institute of Columbia University |  |
|  |  | On Being a Mentor:  A Guide for Higher Education Faculty [6] | Respondent | W. Brad Johnson | A book that offers strategies for effective mentorship |
| Individual | Public Actions | [Men in Global Health, Time to 'Lean Out'](https://communities.springernature.com/posts/men-in-global-health-time-to-lean-out) [7] | Research team | Madhukar Pai | An article that outlines actions men can take to support women in global health |
| Individual | Meetings and events | [Event Organizer's Checklist](https://media.wix.com/ugd/ffa4bc_21c432e3226c46ca88c29b50ba64996b.pdf) [8] | Research team | Women Leaders in Global Health Initiative | A checklist of key points for event organizers to ensure gender equity |
| Individual | Meetings and partnerships | [Equitable and Efficient Collaborations Toolkit](https://ctb.github.io/2023-equitable-efficient/) [9] | Respondent | University of California Davis | A toolkit for organizing and facilitating meetings and collaborations to improve equity and efficiency |
| Individual | Training | [Unconscious Bias in Medicine](https://online.stanford.edu/courses/som-ycme0027-unconscious-bias-medicine) [10] | Research team | Stanford Online and EdX | An online course on recognizing unconscious bias and the impact on decision making |
| Individual | Multitude of actions | [How Male Allies Can Support the Advancement of Women in Academic Medicine](https://journals.lww.com/academicmedicine/fulltext/2023/08000/how_male_allies_can_support_the_advancement_of.31.aspx) [11] | Research team | Sinha et al. | An infographic of 10 strategies to help men be better allies |
|  |  | Good Guys: How Men Can Be Better Allies for Women in the Workplace [12] | Research team | David G. Smith and W. Brad Johnson | A book to help guide men to be better allies for their women colleagues |
|  |  | Better Allies: Everyday Actions to Create Inclusive, Engaging Workplaces [13] | Research team | Karen Catlin | A book on cultivating allyship in the workplace |
|  |  | [7 Tips for Men Who Want to Support Equality](https://leanin.org/tips/mvp#!) [14] | Research team | Lean In | A list of tips for men to support gender equity |
| Institutional | Policies and programs | [Male Allyship Toolkit](https://www.heforshe.org/en/heforshe-alliance-launches-male-allyship-toolkit-resource-building-inclusive-workplaces) [15] | Research team | HeforShe Alliance | A toolkit with case studies encouraging male allyship on an institutional level through programs and policies |
| Institutional | Raise awareness | [Act One: There Will Be Blood](https://www.thisamericanlife.org/840/how-are-you-not-seeing-this/act-one-9) [16] | Research team | This American Life | A podcast episode that can increase awareness and accommodations for biological differences between men and women |
| Institutional | Training | [StanfordOnline: Strategies to Improve and Incorporate Diversity, Equity and Inclusion in Healthcare](https://www.edx.org/learn/medicine/stanford-university-strategies-to-improve-and-incorporate-dei-in-healthcare) [17] | Research team | edX and Stanford Online | An online open-access course offering best practices for leaders to build and sustain teams to enhance equity |
| Societal | Supporting young boys to be better allies | BoyMom: Reimagining Boyhood in the Age of Impossible Masculinity [18] | Research team | Ruth Whippman | A book on understanding the current culture impacting boyhood and strategies to navigate raising boys in such an environment |
|  |  | Raising Feminist Boys: How to Talk with Your Child about Gender, Consent, and Empathy [19] | Research team | Bobbi Wegner and Jessica Joelle Alexander | A book offering parents guidance to discuss gender, empathy, identity and other related topics |
| Societal | Shared caregiving | [Fair Play](https://www.fairplaylife.com/) [20,21] | Research team | Book: Eve Rodsky  Movie: Jennifer Siebel Newsom and Hello Sunshine | A book and documentary film offering a system for shared parenting and equal responsibilities in caregiving |
| Societal | Advocacy | [Equimundo](https://www.equimundo.org/),  [MenCare](https://www.mencare.org/resources/state-of-americas-fathers/),  [Global Boyhood Initiative](https://www.boyhoodinitiative.org/) [22–24] | Research team |  | A collective that promotes allyship amongst men and boys and promotes men as caregivers |

1.	What Every Health Researcher Needs to Know About Health Equity: Privilege, Oppression and Allyship [Internet]. 2019 [cited 2025 May 12]. Available from: https://www.youtube.com/watch?v=yiWZK2AxF7M

2.	Guidance for Avoiding All Male Panels [Internet]. United Nations Indonesia; Available from: https://indonesia.un.org/sites/default/files/2021-06/Guidance_No%20Manel_0.pdf

3.	Mentoring Checklists [Internet]. The Center for Faculty Excellence at the GW School of Medicine and Health Sciences. [cited 2025 May 12]. Available from: https://cfe.smhs.gwu.edu/mentoring-checklists

4.	Avoiding gender bias in reference writing. Commission on the Status of Women, University of Arizona;

5.	Dutt K. Avoid Implicit Gender Bias in Recommendation Letters. Lamont-Doherty Earth Observatory, The Earth Institute of Columbia University; 2019.

6.	Johnson WB. On being a mentor: a guide for higher education faculty. Second Edition. New York: Routledge; 2016. 304 p.

12.	Smith DG, Johnson WB. Good Guys: how men can be better allies for women in the workplace. Boston (Mass.): Harvard business review press; 2020.

13.	Catlin K. Better Allies: Everyday Actions to Create Inclusive, Engaging Workplaces. 2nd ed. Karen Catlin Consulting; 2019. 253 p.

14.	7 Tips for Men Who Want to Support Equality [Internet]. Lean In. [cited 2025 May 14]. Available from: https://leanin.org/tips/mvp

15.	Male Allyship Toolkit [Internet]. HeForShe Alliance; 2024 [cited 2025 May 13]. Available from: https://www.heforshe.org/en/heforshe-alliance-launches-male-allyship-toolkit-resource-building-inclusive-workplaces

16.	There Will Be Blood [Internet]. This American Life. 2024 [cited 2025 May 15]. Available from: https://www.thisamericanlife.org/840/how-are-you-not-seeing-this/act-one-9

17.	Maldonado Y, Fassiotto M. StanfordOnline: Strategies to Improve and Incorporate Diversity, Equity and Inclusion in Healthcare [Internet]. edX. [cited 2025 Aug 7]. Available from: https://www.edx.org/learn/medicine/stanford-university-strategies-to-improve-and-incorporate-dei-in-healthcare

18.	Whippman R. BoyMom: Reimagining Boyhood in the Age of Impossible Masculinity. Harmony; 2024. 320 p.

19.	Wegner B, Alexander JJ. Raising feminist boys: how to talk with your child about gender, consent & empathy. Oakland: New Harbinger Publications; 2021. 1 p.
